# Development of Machine Learning-Based Mpox Surveillance Models in a Learning Health System

**DOI:** 10.1101/2024.09.25.24314318

**Authors:** Harry Reyes Nieva, Jason Zucker, Emma Tucker, Jacob McLean, Clare DeLaurentis, Shauna Gunaratne, Noémie Elhadad

## Abstract

We developed machine learning and deep learning models to identify mpox cases from clinical notes as part of a learning health system initiative. Lasso regression outperformed deep learning models, excelled in minimizing false positives, and may prove helpful for flagging missed or delayed diagnoses as part of continuous quality improvement.

## BACKGROUND

In 2022, outbreaks of mpox (formerly known as monkeypox) spread across Europe, the Americas, and Australia. During that year, there were 3,829 mpox cases in New York City (NYC).^1^ Mpox symptoms can include painful rash, lymphadenopathy, fever, fatigue, myalgia, headache, and respiratory symptoms. Illness typically ranges from two to four weeks. Despite substantial reduction of cases in NYC following roll-out of the JYNNEOS vaccine, there are still sporadic outbreaks in cities across the United States.^2^ In August 2024, the World Health Organization declared a public health emergency in response to new outbreaks emerging from the Democratic Republic of Congo and neighboring countries.^3^ These outbreaks are tied to a new strain belonging to clade I mpox, which is associated with more severe disease than the clade II virus that circulated in 2022.^4^

Efforts to develop machine learning (ML) models that identify mpox at point-of-care have largely focused on utilizing images of skin lesions and deep learning (DL) approaches,^5^ though such models have not been widely deployed. Clinical narratives in the electronic health record (EHR) present a rich and underutilized data source, often containing information (e.g., signs and symptoms) not otherwise found in the clinical record. ML and DL-based classification models that incorporate natural language processing (NLP) have been shown to achieve state-of-the-art performance when applied to other diseases.^6,7^

In addition to point-of-care applications, ML and DL classifiers can also support myriad tasks that enable a learning health system (LHS), such as facilitating clinical quality improvement and assurance efforts via automated case review and stratified population-level assessment of outcomes to spotlight and elucidate potential disparities.^8^ The aim of this study was to develop robust ML and DL-based models capable of detecting mpox cases for surveillance efforts based on content from clinical notes.

## METHODS

### Data, participants, and study setting

The LHS initiative at Columbia University is a collaboration between NewYork-Presbyterian (NYP), Columbia University Irving Medical Center (CUIMC), ColumbiaDoctors, Columbia Engineering, New York State Psychiatric Institute, and the Irving Institute for Clinical and Translational Research, as well as other collaborators across the Columbia campuses. As part of this initiative, we developed mpox classification models based on retrospective study of clinical encounters at NYP, a comprehensive, integrated academic health care delivery system in NYC that sees more than 2 million emergency, primary, and specialty care visits annually.

We restricted our review to clinical visits occurring between May 15, 2022 (at the start of the mpox outbreak) and October 15, 2022 (as mpox rates were in decline due to vaccination). Throughout this period, study team members in the CUIMC Division of Infectious Diseases maintained a prospective list of all patients with mpox diagnoses confirmed by polymerase chain reaction (PCR) testing at the institution and stored this information in a dedicated REDCap database. This study was approved by the CUIMC institutional review board (Protocol AAAU3052).

For each patient with a PCR-confirmed diagnosis, we extracted their electronic health record and administrative data, which was standardized using the Observational Medical Outcomes Partnership Common Data Model.^9^ Our extract included patient demographics and all clinical notes recorded from the date of diagnosis up to 30 days prior to diagnosis. This 30-day look-back window was selected because mpox symptoms typically last between two to four weeks. For each PCR-confirmed case, we also extracted equivalent data for three controls randomly selected by matching on patient age, sex, race, ethnicity, and visit month (to account for secular trends). Due to data limitations, for controls, it is not known if the recorded sex reflected gender identity and/or sex assigned at birth.

### Model development

We trained three mpox classifiers using 70% of the dataset for training and 30% to evaluate model performance. Our three mpox classifiers were developed using 1) logistic regression with L1 regularization, also known as Least Absolute Shrinkage and Selection Operator (LASSO) regression;^10^ 2) ClinicalBERT,^11^ and 3) ClinicalLongformer.^12^

Prior to applying LASSO, we performed several text pre-processing steps that included transforming each word to lowercase, removing punctuation and stopwords (e.g., common terms such as “is”, “the”), and tokenizing text. We also augmented our corpus by generating bi-grams (e.g., “smoking_cessation”) and tri-grams (e.g., “human_immunodeficiency_virus”), then constructed a document-term matrix wherein each term was a potential feature in our model using a count vectorizer.

ClinicalBERT and ClinicalLongformer are pre-trained language models for clinical text. They often achieve state-of-the-art results and can be fine-tuned to perform specific clinical NLP tasks, including classification, through additional training on new data. ClinicalBERT and ClinicalLongformer differ most notably in their ability to model long-term dependencies (i.e., the context of surrounding terms) in long clinical texts. The maximum sequence length for ClinicalBERT is 512 tokens, while ClinicalLongformer can accommodate 4096 tokens. When training each of these DL models, we used a batch size of 32, a learning rate of 5e-5, and trained for 5 epochs.

### Model evaluation

We evaluated model performance using precision (positive predictive value, PPV), recall (sensitivity), F1 score, area under the receiver operating characteristic curve (AUROC), and area under the precision-recall curve (AUPRC). We also computed recall at a precision of 80% (RP80) to minimize false positives and risk of alert fatigue. We performed all model development and evaluation using the Python programming language (Python Software Foundation).

## RESULTS

We identified 228 patients with PCR-confirmed mpox diagnoses during the review period and 698 as controls (**Table 1**). Patient median age was 34 (IQR: 29, 42); recorded sex was male for 902 patients (97%) and female for 24 (3%). Our sample comprised 249 (27%) patients who identified as non-Hispanic Black or African American, 117 (13%) as non-Hispanic white, and 316 (34%) as Hispanic or Latino; 244 (26%) were of unknown race or ethnicity. Based on NYC Department of Health and Mental Hygiene (DOHMH) data, our sample represents 6% of all mpox cases reported in NYC in 2022 and shares a similar demographic composition to citywide cases.^1^

**Table 1:** Cohort Characteristics.

| **Characteristic** | **Overall**<br>**(N=926)** | **Cases**<br>**(N=228)** | **Controls**<br>**(N=698)** |
| --- | --- | --- | --- |
|  | median (IQR) |  |  |
| **Age, years** | 34 (29, 42) | 34 (29, 42) | 34 (29, 42) |
|  | N (%) |  |  |
| **Administrative sex\*** |  |  |  |
| Female | 24 (3%) | 6 (3%) | 18 (3%) |
| Male | 902 (97%) | 222 (97%) | 680 (97%) |
| **Race or ethnicity** |  |  |  |
| Black or African American | 249 (27%) | 60 (26%) | 189 (27%) |
| White | 244 (26%) | 60 (26%) | 184 (26%) |
| Hispanic or Latino | 316 (34%) | 79 (35%) | 237 (34%) |
| Unknown | 117 (13%) | 29 (13%) | 88 (13%) |

\*For matching purposes, we were only able to use available structured data (i.e., administrative sex used for billing purposes, which may not reflect gender identify and/or sex assigned at birth).

For the LASSO regression, precision, recall, and F1 score were all 0.93, the AUROC was 0.97, AUPRC was 0.93, and RP80 was 0.89 (**Table 2, Figure 1**). Phrases related to lesions, pain, genitourinary tenderness, exudate, and HIV were among the most predictive LASSO features aside from mpox mentions (**Table 3**). The ClinicalBERT model achieved a precision of 0.88, recall of 0.89, F1 score of 0.88, AUROC of 0.93, and RP80 of 0.67. The ClinicalLongformer model achieved a precision of 0.87, recall of 0.88, F1 of 0.87, AUROC of 0.92, and RP80 of 0.47.

**Table 2:** Classification Report for Machine Learning and Deep Learning Classifiers for Mpox.

|  | **Precision (PPV)** | **Recall (sensitivity)** | **F1 Score** | **AUROC** | **AUPRC** | **RP80** |
| --- | --- | --- | --- | --- | --- | --- |
| **LASSO Regression** | 0.93 | 0.93 | 0.93 | 0.97 | 0.93 | 0.89 |
| **ClinicalBERT** | 0.88 | 0.89 | 0.88 | 0.93 | 0.82 | 0.67 |
| **ClinicalLongformer** | 0.87 | 0.88 | 0.87 | 0.92 | 0.81 | 0.47 |

*Abbreviations:* AUPRC, area under the precision-recall curve. AUROC, area under the receiver operating characteristic curve. PPV, positive predictive value. RP80, recall at precision of 80%.

**Table 3:** Informative words or phrases and their frequency among mpox cases and controls.

| Word or Phrase | Overall<br>(N=926) | Cases<br>(N=228) | Controls<br>(N=698) |
| --- | --- | --- | --- |
|  | N (%) |  |  |
| abdominal tenderness | 78 (8) | 66 (29) | 12 (2) |
| erythematous | 91 (10) | 46 (20) | 45 (6) |
| exudate | 197 (21) | 92 (40) | 105 (15) |
| HIV, human immunodeficiency virus | 391 (42) | 198 (87) | 193 (28) |
| lesion | 375 (40) | 213 (93) | 162 (23) |
| mpox, mpx, monkeypox, monkey pox | 298 (32) | 212 (93) | 86 (12) |
| nausea and vomiting | 327 (35) | 226 (99) | 101 (14) |
| painful | 207 (22) | 130 (57) | 77 (11) |
| penile lesions | 44 (5) | 41 (18) | 3 (0) |
| positive for rash | 162 (17) | 125 (55) | 37 (5) |
| rectal pain | 144 (16) | 110 (48) | 34 (5) |
| sores | 132 (14) | 105 (46) | 27 (4) |
| strep, streptococcus | 45 (5) | 24 (11) | 21 (3) |
| syphilis | 120 (13) | 101 (44) | 19 (3) |

**Figure 1:**
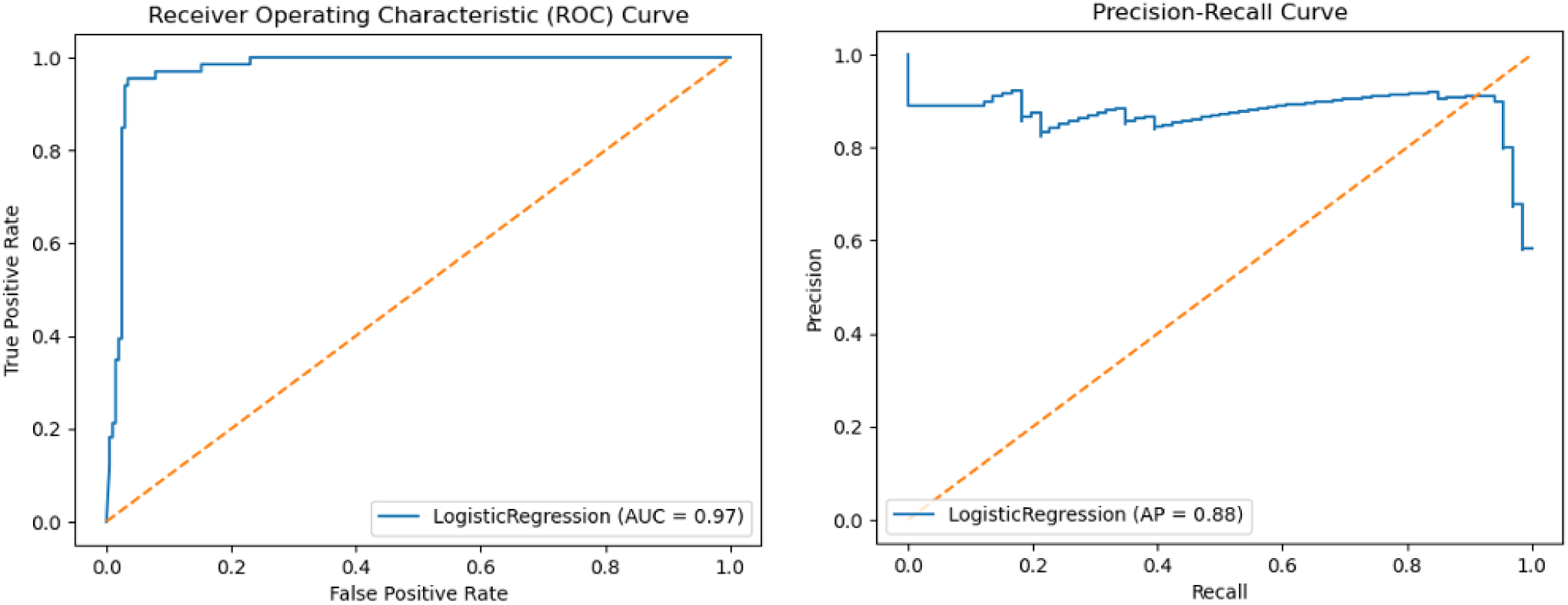
Receiver Operating Characteristic (ROC) and Precision-Recall Curves for LASSO Regression.

## DISCUSSION

As part of an ongoing LHS initiative, we prospectively compiled a list of PCR-confirmed mpox cases, then developed a set of classification models based on this gold standard cohort. In total, our sample comprised a sizable percentage of all mpox cases reported in NYC in 2022 with a representative demographic makeup.

The ML and DL models we trained on clinical notes show promise for enabling accurate identification of mpox cases. LASSO regression outperformed our two deep learning models, though standard performance metrics (precision, recall, F1 score, AUROC, and AUPRC) were robust for all three. Most notably, our models differed considerably when evaluated using RP80, a less common metric intended to minimize false positives. This distinction is especially relevant to the clinical setting where alert fatigue is often a concern.

Despite the ability of ClinicalBERT and ClinicalLongformer to incorporate the context of surrounding terms in their language models, they did not outperform LASSO regression. Symptoms documented in clinical notes were among the most predictive features in the LASSO regression, which suggests a potential explanation for this finding. First, mere mention of certain symptoms (e.g., lesion) may provide sufficient information for the model to learn, so the context mapped by the language models were not necessarily essential to this particular use case. Second, symptoms are primarily mentioned early in notes, which may render the distinguishing characteristic between the two DL models (i.e., differences in text sequence length) less relevant.

The LHS paradigm involves continuous aggregation and analysis of data and incorporating what is learned into the improvement of future care as part of a natural feedback loop. Operationally, our vision for this process moving forward encompasses re-training and testing models at regular time intervals and applying the best performing model to clinical encounters not originally included in the development process. Such models, for example, would facilitate retrospective case review by flagging missed or delayed mpox diagnoses and inform continuous quality improvement and assurance efforts.

There are several important limitations to consider. First, our models only use text from clinical notes. Future work incorporating structured data may result in improved performance. Second, given that documentation practices may change during and after an outbreak, further study of differences in model performance based on the dates of clinical encounters may also be especially critical to account for model drift.

Our findings illustrate how ML and DL methods may help harness relatively untapped resources (e.g., clinical text) in support of case surveillance and identification. Importantly, application of these methods extends beyond just mpox, potentially providing clinicians and public health officials alike with a valuable tool to combat any infectious disease.

## Acknowledgements

Preliminary data from this study were presented at the Symposium on Artificial Intelligence in Learning Health Systems in Rio Grande, Puerto Rico on May 10, 2023; STI & HIV 2023 World Congress in Chicago, IL on July 25, 2023; and the American Medical Informatics Association 2023 Annual Symposium in New Orleans, LA on November 13, 2023.

## Contributor Statement

All authors had full access to the data in the study. They also take responsibility for the integrity of the data and accuracy of the analysis, and have approved the final manuscript.

*Concept and design:* Reyes Nieva, Elhadad, Zucker

*Acquisition, analysis, or interpretation of data:* All authors

*Drafting of the manuscript:* Reyes Nieva

*Critical revision of the manuscript for important intellectual content:* All authors

*Statistical analysis:* Reyes Nieva

*Obtained funding:* Elhadad, Zucker

*Administrative, technical, or material support:* Reyes Nieva

*Supervision:* Elhadad, Zucker

## Data Availability Statement

The electronic health record and administrative data underlying this article are not available due to restrictions to preserve patient confidentiality.

